# A Task-Optimized Approach for High-Accuracy Alzheimer’s Diagnosis from Handwriting Data

**DOI:** 10.1101/2024.12.17.24319146

**Authors:** Zahra Seyedi HosseiniNian, Ahmadreza Tajari, Behrouz Barati B, Sina Baharlouei

**Affiliations:** Department of Electrical Engineering, Shahrood University of Technology, Shahrood, Iran; Electrical Engineering Department, Sharif University of Technology, Tehran, Iran; Department of Computer Science, California State University, Northridge, 91330, CA, USA; eBay Search Ranking and Monetization, San Jose, 95126, CA, USA

**Keywords:** XGBoost Classifier, Feature Selection, Analysis of Handwriting, Alzheimer’s Disease Diagnosis

## Abstract

Training complex models on Alzheimer’s Disease (AD) datasets is challenging due to the costly process of extracting features from a wide range of patient tasks. Developing high-performance AD detection models that rely on a small number of tasks can help reduce dataset acquisition costs and improve the interpretability of the AD detection model. To address this, we propose a two-stage forward-backward feature selection approach to identify the most relevant tasks and features for predicting AD with high accuracy. We evaluate a range of machine learning methods, including Extreme Gradient Boosting (XGBoost), Random Forest, K-Nearest Neighbors, Support Vector Machine, Multi-Layer Perceptron, and Logistic Regression, to determine the best classification model for feature selection and downstream prediction tasks. Given the limited sample size, we assess model performance using Leave-One-Out-Cross-Validation (LOOCV) to ensure robust results. Our method was compared with multiple state-of-the-art approaches for feature selection. The results of our analysis indicate that combining our proposed methods for feature selection with the XGBoost classifier, using only four tasks, produces a model that is both more interpretable and high-performing compared to other approaches. This suggests focusing on these four tasks, rather than collecting extensive task data from patients, can yield a reliable predictor for diagnosis of AD with an accuracy of 91.37%, 93.94% recall, 89.77% precision, and 91.32% F1 score - surpassing other classification methods. This research represents a significant advancement in the efficiency and reliability of AD diagnosis, improving patient prognosis and offering potential benefits to healthcare systems.

## 1 Introduction

Dementia presents a critical global health challenge, affecting over 55 million people worldwide across diverse socioeconomic landscapes. It disproportionately impacts low- and middle-income countries, where more than 60% of those affected live, and reports indicate nearly 10 million new cases each year. As the seventh leading cause of death globally, dementia not only increases mortality but also significantly contributes to disability and dependency, particularly among the elderly. The economic impact is profound; in 2019, the cost of dementia surpassed 1.3 trillion US dollars. Neurodegenerative disorders, especially AD, account for the majority of dementia cases, representing approximately 60–70% of all diagnoses [30]. AD is characterized by a gradual and relentless decline in cognitive functions, affecting critical areas such as memory, reasoning, judgment, and learning. AD is predominantly marked by episodic memory impairments in its early stages, a hallmark of ventromedial temporal lobe dysfunction [4]. As the disease progresses, these memory deficits worsen into profound amnesia, accompanied by impairments in other cognitive domains, reflecting the widespread pathological involvement of neural networks. This progression highlights the intricate neurobiology of AD and the considerable challenges faced in its early diagnosis and intervention [12]. Once the disease advances to the dementia stage, it remains medically incurable, with current pharmacological treatments only able to slow its progression [28]. This stark prognosis underscores the critical importance of early screening and detection, as identifying AD in its nascent stages offers the best opportunity for timely intervention and potentially slows the disease’s trajectory [12]. Writing is a complex human activity that relies on a sophisticated combination of cognitive, kinesthetic, and perceptual-motor components, including visual and kinesthetic perception, motor planning, eye-hand coordination, visuomotor integration, talent, and manual skills [24]. The brain plays the most critical role in writing, as it learns and develops this ability, which can be used throughout life. However, writing is one of the first abilities to be affected by AD as the brain’s cognitive function declines [36, 45]. Early symptoms of AD, such as memory disorders, difficulties in problem-solving, and decreased responsiveness to daily events, can lead to changes in handwriting and signature. These changes are often subtle initially, becoming more pronounced as the disease progresses. Affected individuals may struggle to recall the text they intended to write or remember letters and signature forms, often requiring a model to guide their writing. In the later stages of AD, the swift and automatic nature of handwriting, particularly in signatures, diminishes, leading to slower, interrupted movements. Further progression of the disease can result in forgetting letters, unnecessary repetition, and illogical connection lines within the text, eventually rendering the handwriting illegible. Handwriting impairments often begin before AD is diagnosed clinically, serving as an essential indicator of cognitive decline [25, 35]. Given these changes, handwriting analysis has emerged as a promising biomarker for assessing AD and other cognitive disorders, providing a basis for early diagnosis through noticeable differences in writing [6, 37].

In AD research, prolonged evaluation protocols can worsen cognitive overload, leading to decreased attention and impaired memory—already compromised in AD patients [14]. Minimizing tasks reduces fatigue, which affects the accuracy of assessments. Fatigue diminishes performance and exacerbates confusion and agitation, leading to unreliable results [2, 46]. Motivated by these points, we aim to find the best subset of tasks where the performance of the trained model on them is as good as the original data. In other words, the key research question relies on what subset of tasks one can predict whether someone has AD with high accuracy compared to the case we consider all possible tasks. This can help researchers collect data on those specific tasks instead of many other tasks that do not add significant prediction power. To address this, we propose a two-stage forward-backward feature selection approach to identify the most relevant tasks and features for predicting AD with high accuracy. Shorter, targeted assessments improve patient compliance and provide more accurate reflections of cognitive abilities. Streamlined protocols save time and costs for healthcare providers and patients [3, 23, 27, 31]. Reduced testing times allow clinicians to evaluate more patients, decreasing operational costs and improving resource allocation. For patients, shorter assessments mean less disruption and potentially lower expenses [13, 15, 16, 23, 32, 33]. Concise assessments also alleviate anxiety and discomfort associated with longer testing. Our approach enhances comfort and encourages participation by enabling quicker completion and promoting accurate and frequent monitoring. This patient-friendly approach fosters a positive patient-clinician relationship and supports AD management while adhering to ethical guidelines emphasizing patient autonomy, beneficence, and non-maleficence. By minimizing potential harm and maximizing welfare, our methodology upholds the dignity of individuals while optimizing diagnostic accuracy and aligning with best practices for patient-centered care [1, 8, 18, 40, 42]. Therefore, our methodology optimizes diagnostic accuracy and aligns with best practices for patient-centered care in neuropsychological evaluations.

### 1.1 Related Works

In recent years, the application of machine learning in healthcare has grown significantly, helping to address various challenges and advance the field [5, 19, 21]. Machine learning models have become increasingly crucial as computer-assisted systems for diagnosing neurodegenerative diseases [44].

Researchers developed diverse ensemble models analyzing handwriting kinetics by employing a stacking technique to combine multiple base-level classifiers. In particular, Cilia et al. [11] published a study that included 174 participants, including 89 individuals diagnosed with AD and 85 healthy individuals, sourced from the DARWIN dataset [11]. Önder et al. [34] in a thorough investigation aimed at diagnosing AD applied and compared four distinct classification techniques: XGBoost, GradientBoost, AdaBoost, and voting classifiers. Among these methods, XGBoost emerged as the most accurate, achieving an 85% accuracy rate for AD diagnosis.

Gattulli et al. [17] utilized machine learning-based classifiers with high-performance scores to solve the difficulty of manual AD detection. In this study, Light Gradient Boosting Machine (LightGBM), Categorical Boosting (CatBoost), and Adaptive Boosting (AdaBoost) machine learning classification algorithms were combined with a Hard Voting Classifier and trained and tested on the DARWIN dataset. As a result, the proposed Ensemble methodology achieved 97.14% Acc, 95% Precision, 100% Recall, 90.25% Spec, and 97.44% F1-score performance values.

There is evidence suggesting that not all tasks are equally important in assessing a patient’s health status. Subha et al. [43] proposed a selection of handwriting tasks based on an analysis of challenging cases within the DARWIN dataset, which contains 25 online-recorded handwriting tasks, each characterized by a standard set of features. Several classification models—including Random Forest (RF), Logistic Regression (LR), K-Nearest Neighbor (KNN), Linear Discriminant Analysis (LDA), Support Vector Machines (SVM), Bayesian Networks (BN), Gaussian Naïve Bayes (GNB), Multilayer Perceptron (MP), and Learning Vector Quantization (LVQ)—were used to identify users who were frequently misclassified. The selected tasks varied in terms of writing and drawing types and their complexity levels. The methodology for selecting handwriting tasks relies on statistical and similarity analysis of classification results to identify patterns in misclassification. Experiments were repeated 20 times to ensure reliability, and the classifiers were optimized using grid search with 5-fold cross-validation to determine the best hyperparameters. The experiments were conducted on the entire dataset and the selected task group, revealing that some patients with AD were misclassified as healthy at least once by all classifiers, underscoring the presence of commonly misclassified individuals.

A swarm intelligence-based feature selection approach was combined with several machine learning models, including Logistic Regression (LR), K-Nearest Neighbors (KNN), Support Vector Machine (SVM), Decision Tree (DT), Random Forest (RF), and AdaBoost, to develop a hybrid Machine Learning (ML) model for AD detection. The RF and AdaBoost classifiers achieved the highest performance, yielding an accuracy of 90%, precision of 88%, recall of 92%, F1-score of 90%, and an Area Under the Receiver Operating Characteristic Curve (AUC-ROC) score of 90% [43]. In another study, Mitra and Rehman [30] adopts different feature selection techniques to enhance the model’s interpretability and a low number of data points relative to the number of features. Both Repeated k-fold and Monte-Carlo cross-validation techniques were applied to evaluate the model. Furthermore, the Analysis of Variance (ANOVA) was utilized to select the top *k* features for each best-performing base classifier. The results achieved 97.14% accuracy, 95% sensitivity, 100% specificity, 100% precision, 97.44% F1-score, 94.37% Matthews Correlation Coefficient (MCC), 94.21% Cohen Kappa, and 97.5% AUC-ROC [30].

A novel particle swarm optimization algorithm has been developed to fine-tune hyperparameters within Convolutional Neural Network (CNN) architectures, significantly improving classification accuracy for AD severity [26]. This model achieved an impressive 99.53% accuracy and 99.63% F1-score on a public dataset, which performs better than previous studies [26]. The enhanced model could streamline doctors’ decision-making processes.

Hakan [22] investigates AD by developing a machine learning model that uses handwriting data to classify individuals as AD patients. Principal Component Analysis (PCA) is employed as a data preprocessing technique. At the same time, the model training involves a comparison of a linear support vector classifier, a random forest classifier, and XGBoost, along with their respective accuracy metrics. Each participant’s handwriting sample produces 451 features, which serve as the input parameters for the model. The results indicate that when applied with PCA to reduce the feature set to 65, the Random Forest classifier achieved the highest accuracy at 94.29%. Future implications and potential impact of this promising advancement in AI-driven AD diagnosis may involve utilizing more extensive and diverse datasets to further improve model generalizability Kaya and Çetin-Kaya [26].

### 1.2 Motivation and Contributions

While many machine learning pipelines have been proposed to maximize the accuracy of AD detection, the interpretability of these models is often overlooked. This paper aims not to develop complex ensemble models that achieve near-perfect performance on small datasets (fewer than 500 samples). First and foremost, the generalizability of such models to other AD-related datasets cannot be assumed based solely on good performance with fewer than 500 data points. Moreover, evaluation metrics like random train-test splits and *K*-fold cross-validation with small *k* are unreliable, as demonstrated by our experiments, which show significant performance variance. Finally, achieving near-perfect accuracy without interpreting the results or identifying the most important features contributing to the model’s performance may not give researchers the insights to understand the critical factors influencing the prediction of AD.

Motivated by these considerations, our goal is to identify the optimal subset of tasks for which the performance of the trained model is comparable to that achieved using the complete set of tasks. In other words, the central research question is: What subset of tasks allows for high-accuracy AD prediction compared to using all available tasks? This approach can guide researchers in focusing data collection efforts on the most predictive tasks rather than tasks that do not contribute significantly to the model’s accuracy.

To achieve the aforementioned goal, this paper presents a machine learning pipeline (see Figure 1) that provides an interpretable Alzheimer’s disease (AD) detector, using only 50% of the available tasks and less than 10% of the features. Task and feature selection are performed in two sequential stages, employing a forward-backward feature selection technique with XGBoost as the base classifier. By extracting relevant tasks and features and applying the XGBoost classifier, we aim to enhance diagnostic accuracy and efficiency. Furthermore, the ability to rely on fewer tasks without sacrificing model performance suggests which tasks are most important for detecting Alzheimer’s, providing valuable insights for future data collection efforts. Finally, we conduct a comparative analysis of our model with various machine learning methods, each utilizing different feature selection strategies.

**Fig 1.**
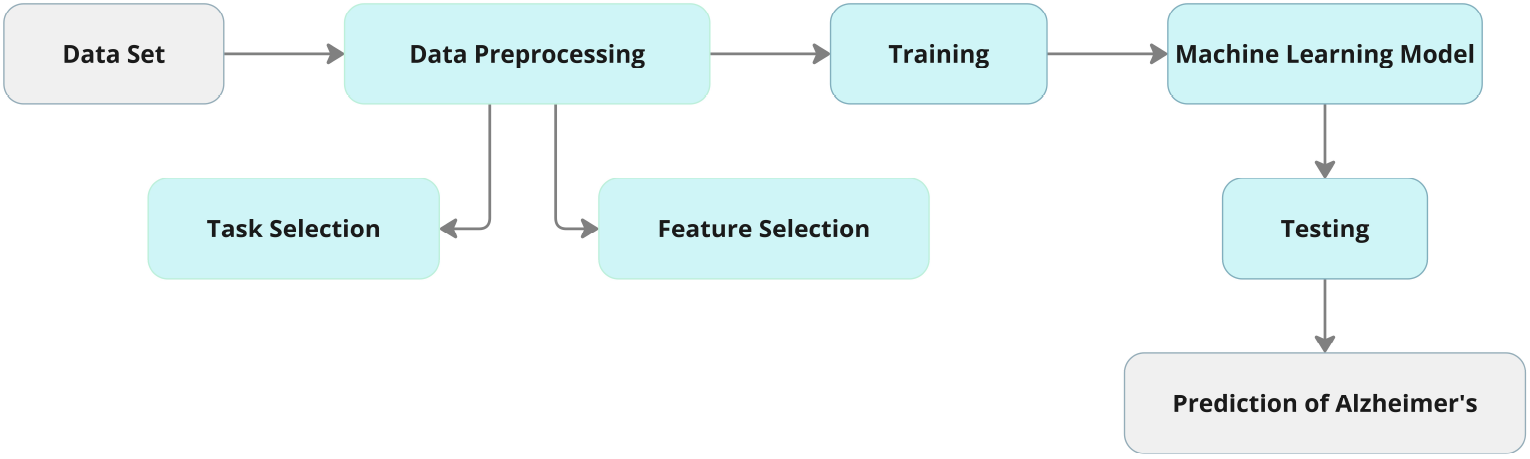
The methodology pipeline including, pre-processing (task and feature selection), training, model selection (hyper-parameters), and model evaluation (testing)

This work is primarily characterized by:

1. Proposing a two-step forward-backward feature selection technique to pinpoint the most relevant tasks and features contributing to AD detection to improve diagnostic efficiency, particularly with small training samples.
2. Advancing AI-based diagnostic methods by integrating XGBoost with the proposed feature selection approach, ultimately creating tools that are more accurate, accessible, and cost-effective to improve patient outcomes and lower healthcare expenses.

The rest of the article is organized as follows. Section 2 outlines the methodology, including data preparation, the XGboost structure, feature selection techniques, and evaluation criteria. Section 3 focuses on implementing feature extraction and evaluating XGBoost’s performance compared to other classifiers. The results show that the XGBoost with forward-backward feature selection outperformed other models, even with a small training sample size. Section 4 presents the article’s conclusion and summarizes the essential findings.

## 2 Methods

In this section, we first provide a detailed description of the DARWIN (Diagnosis Alzheimer With Handwriting) dataset (Section 2.1). Then, we outline the methodology pipeline, which includes the XGBoost predictor, the task and feature selection algorithm, and the evaluation strategy used to identify the final model (Section 2.2 and Section 2.3).

### 2.1 Dataset Description

The DARWIN (Diagnosis Alzheimer With Handwriting) dataset utilized in this study comprises handwriting data collected from 174 participants, including 89 AD patients and 85 healthy controls, as described in Cilia et al. [11]. The dataset contains hand-writing samples obtained from 25 distinct tasks designed to assess various aspects of motor and cognitive function. These tasks are divided into four categories: graphic tasks, copy tasks, memory tasks, and dictation tasks (see Table 1). The tasks included activities such as drawing geometric figures, copying letters and words, writing under dictation, and recalling memorized items. Each task was intended to evaluate different dimensions of handwriting abilities and their association with cognitive function. Participants were recruited based on standard clinical assessments—Mini-Mental State Examination (MMSE), Frontal Assessment Battery (FAB), and Montreal Cognitive Assessment (MoCA)—to ensure a comprehensive evaluation of their cognitive abilities. The dataset includes the pen’s on-paper and in-air movements, recorded using a Wacom Bamboo tablet at a frequency of 200 Hz, along with pressure data during on-paper writing. To maintain demographic consistency and minimize bias, participants from both groups were matched based on age, education level, type of work, and gender. Data acquisition involved using a standardized protocol in which participants wrote on A4 paper sheets placed on a tablet. The collected data were subsequently processed to extract 18 features from each task, capturing fine motor and cognitive variations relevant to distinguishing AD patients from healthy controls (see Table 2). These features provide insights into the subtle changes in handwriting associated with cognitive decline, as documented in the DARWIN dataset [12].

**Table 1.** Overview of handwriting tasks performed, categorized as memory and dictation (M), graphic (G), or copy (C).

| # | Description | Category |
| --- | --- | --- |
| 1 | Signature drawing | M |
| 2 | Join two points with a horizontal line, continuously for four times | G |
| 3 | Join two points with a vertical line, continuously for four times | G |
| 4 | Retrace a circle (6 cm of diameter) continuously for four times | G |
| 5 | Retrace a circle (3 cm of diameter) continuously for four times | G |
| 6 | Copy the letters 'l', 'm', and 'p' | C |
| 7 | Copy the letters on the adjacent rows | C |
| 8 | Write cursively a sequence of four lowercase letter 'l', in a single smooth movement | C |
| 9 | Write cursively a sequence of four lowercase cursive bigram 'le', in a single smooth movement | C |
| 10 | Copy the word 'foglio' | C |
| 11 | Copy the word 'foglio' above a line | C |
| 12 | Copy the word 'mamma' | C |
| 13 | Copy the word 'mamma' above a line | C |
| 14 | Memorize the words 'telefono', 'cane', and 'negozio' and rewrite them | M |
| 15 | Copy in reverse the word 'bottiglia' | C |
| 16 | Copy in reverse the word 'casa' | C |
| 17 | Copy six words (regular, non-regular, non-words) in the appropriate boxes | C |
| 18 | Write the name of the object shown in a picture (a chair) | M |
| 19 | Copy the fields of a postal order | C |
| 20 | Write a simple sentence under dictation | M |
| 21 | Retrace a complex form | G |
| 22 | Copy a telephone number | C |
| 23 | Write a telephone number under dictation | M |
| 24 | Draw a clock, with all hours and put hands at 11:05 (Clock Drawing Test) | G |
| 25 | Copy a paragraph | C |

**Table 2.** Overview of features extracted from handwriting tasks, providing details on motor and cognitive aspects measured.

| Feature | Description |
| --- | --- |
| Total Time (TT) | Total time spent to perform the entire task. |
| Air Time (AT) | Time spent to perform in-air movements. |
| Paper Time (PT) | Time spent to perform on-paper movements. |
| Mean Speed on-paper (MSP) | Average speed of on-paper movements. Speed is the variation of displacement with respect to time. |
| Mean Speed in-air (MSA) | Average speed of in-air movements. |
| Mean Acceleration on-paper (MAP) | Average acceleration of on-paper movements. Acceleration is the variation of speed with respect to time. |
| Mean Acceleration in-air (MAA) | Average acceleration of in-air movements. |
| Mean Jerk on-paper (MJP) | Average jerk of on-paper movements. Jerk is the variation of acceleration with respect to time. |
| Mean Jerk in-air (MJA) | Average jerk of in-air movements. |
| Pressure Mean (PM) | Average of the pressure levels exerted by the pen tip. |
| Pressure Var (PV) | Variance of the pressure levels exerted by the pen tip. |
| GMRT on-paper (GM RTP) | Generalization of the Mean Relative Tremor (MRT) computed for on-paper movements. |
| GMRT in-air (GM RTA) | Generalization of the Mean Relative Tremor computed for in-air movements. |
| Mean GMRT (GMRT) | Average of GM RTP and GM RTA. |
| Pendowns Number (PWN) | Counts the total number of pendowns recorded during the execution of the entire task. |
| Max X Extension (XE) | Maximum extension recorded along the X axis. |
| Max Y Extension (YE) | Maximum extension recorded along the Y axis. |
| Dispersion Index (DI) | Measures how the handwritten trace is 'dispersed' on the entire piece of paper. |

### 2.2 Classifier Description and Evaluation Strategy

We utilize XGBoost [10] classifier to detect Alzheimer’s disease in patients based on 450 features from the DARWIN dataset. XGBoost builds an ensemble of decision trees in a sequential, greedy manner. In each iteration, a new tree is trained to fit the negative gradient of the loss function. Intuitively, this means that each new tree is focused on correcting the errors made by previous trees, particularly for data points where the loss is highest. The final model is a weighted combination of all the individual trees.

Mathematically speaking, let 𝒟_train_ = {(**x**_1_, *y*_1_), …, (**x**_*n*_, *y*_*n*_)} be *n* training data points, and 𝓁 is the loss function (e.g., mean squared loss). The objective is to optimize the following model over *n* training data points:

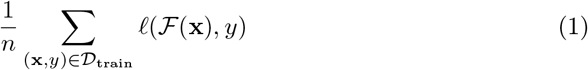

The model is initialized with a best constant predictor of the loss function over *n* data points:

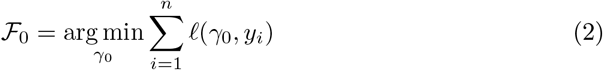

Then, *M* trees will be learned additively in the following manner. At iteration 1 ≤ *m* ≤ *M*, we first compute the pseudo-residual errors of *n* data points:

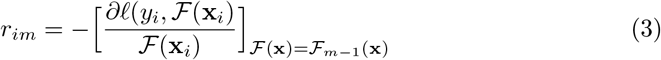

Then, a new tree 𝒯_*m*_ is trained on the computed pseudo-residuals. Next, the optimal coefficient of the new tree is computed by solving a single-parameter optimization problem as follows:

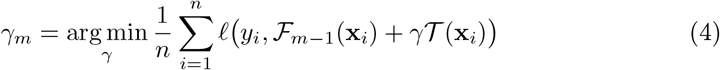

The final predictor in stage *m* will be:

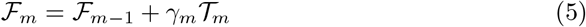

To select the best possible set of hyper-parameters to maximize the performance, we apply the **Leave-One-Out** approach (K-fold cross-validation when *K* = *n*, the number of data points) [47]. In particular, Leave-One-Out leaves one of the data points for validation and *n* − 1 for training. The training model will be evaluated on a single data point. This procedure is repeated on all *n* data points (therefore, *n* models must be trained).

Leave-One-Out as the evaluation metric is more reliable than other methods, such as randomly separating training and test data and *K*-fold cross-validation with small *K* [41]. In particular, it is a perfect choice for the model evaluation on datasets with few data points, such as the DARWIN dataset [29]. The main issue of the Leave-One-Out approach is that it is computationally expensive compared to the *K*-fold cross-validation with small *K* (*K* ≤ 10) [9]. However, since the DARWIN dataset is not large-scale (the number of samples is less than 1000), the training time does not exceed a few minutes using Leave-One-Out as the evaluation metric. Furthermore, our experiments show that the performance reports of many other papers in the literature highly depend on their evaluation method. Therefore, multiple runs of their methods show a very high variance in terms of performance if *K* fold cross-validation or random separation of training and test data is adopted. The main reason is that AD data have a small number of samples, and evaluating performance on a small number of data points might not be reliable. Therefore, we recommend using the Leave-One-Out, which is feasible for this specific dataset without using approximation techniques such as Jackknife estimator [20].

We performed a grid search over several hyperparameters: the number of trees ({10, 20, 30, 50, 100, 200, 500, 1000}), maximum depth of the trees ({2, …, 7}), learning rate ({0.01, 0.02, 0.05, 0.1, 0.2, 0.5}), fraction of features used by each tree ({0.7, 0.8, 0.9, 1}), and fraction of used samples for each tree ({0.8, 0.9, 1}). This search aimed to identify the optimal combination of hyperparameters that maximizes performance, as measured by Leave-One-Out cross-validation accuracy. The resulting best combination of hyperparameters was 50 trees, a maximum depth of 4, a learning rate of 0.3, a feature fraction of 0.8, and a sample fraction of 1.

### 2.3 Task and Feature Selection Algorithm

This section describes the methodology of selecting the optimal set of tasks and features to minimize the number of tasks and the utilized features while maintaining the model’s performance. In the first stage, we choose a subset of tasks demonstrating the highest performance in terms of AUC measured by the Leave-One-Out evaluation metric. The idea is to initialize the task pool with all 8 tasks. Then, at each iteration, the task that increases the performance of the XGBoost model by the most in terms of Area Under Curve (AUC) will be selected and added to the final set of selected tasks. This procedure requires 𝒪(*T* ^2^) (*T* is the number of tasks) model training. To reduce the training time, a simple but effective heuristic inspired by [7] is to remove the tasks, not increasing the performance from the task pool at each iteration of the procedure. Finally, a backward step will be applied if eliminating one of the selected tasks does not increase the performance in the final model. The procedure is presented in Algorithm 1. After choosing the best subset of tasks, the second stage will be applied to all features within the selected tasks. In other words, using these two stages, we simplify the exhaustive method of forward-backward selection. As a result, instead of 𝒪 (*d*^2^) model training that requires more than 24 hours for *d >* 400 (given the Leave-One-Out evaluation procedure), we first choose the best subset of tasks among those 8. Then, we further refine the feature set by applying feature selection to those tasks’ features.

After running the algorithm on the task pool, we apply backward feature selection to the model. However, our experiments show that the selected tasks remain unchanged after the backward pass. This indicates that applying Algorithm 1 alone is sufficient for task selection.

One key advantage of this task and feature selection approach is that, unlike ANOVA-based methods, it is not restricted to linear correlations. As shown in the literature, linear models often fail to capture the complex relationships between features and the target variable, particularly in the healthcare domain [38, 39]. Furthermore, calculating pairwise correlations between each feature and the target variable overlooks the intricate, higher-order interactions among the features. A potential drawback of this approach is its computational expense. However, this issue is addressed through two key strategies. First, instead of applying the method to individual features (450 features), we group them based on the tasks performed by the patients (8 tasks, each with 25 features). Additionally, underperforming features are eliminated from the candidate list using the pruning technique described earlier. Our results show that, on average, nearly 10% of features are removed in each outer loop iteration, significantly reducing the total number of iterations required.

#### Algorithm 1

Forward Task (Feature) Selection with Pruning

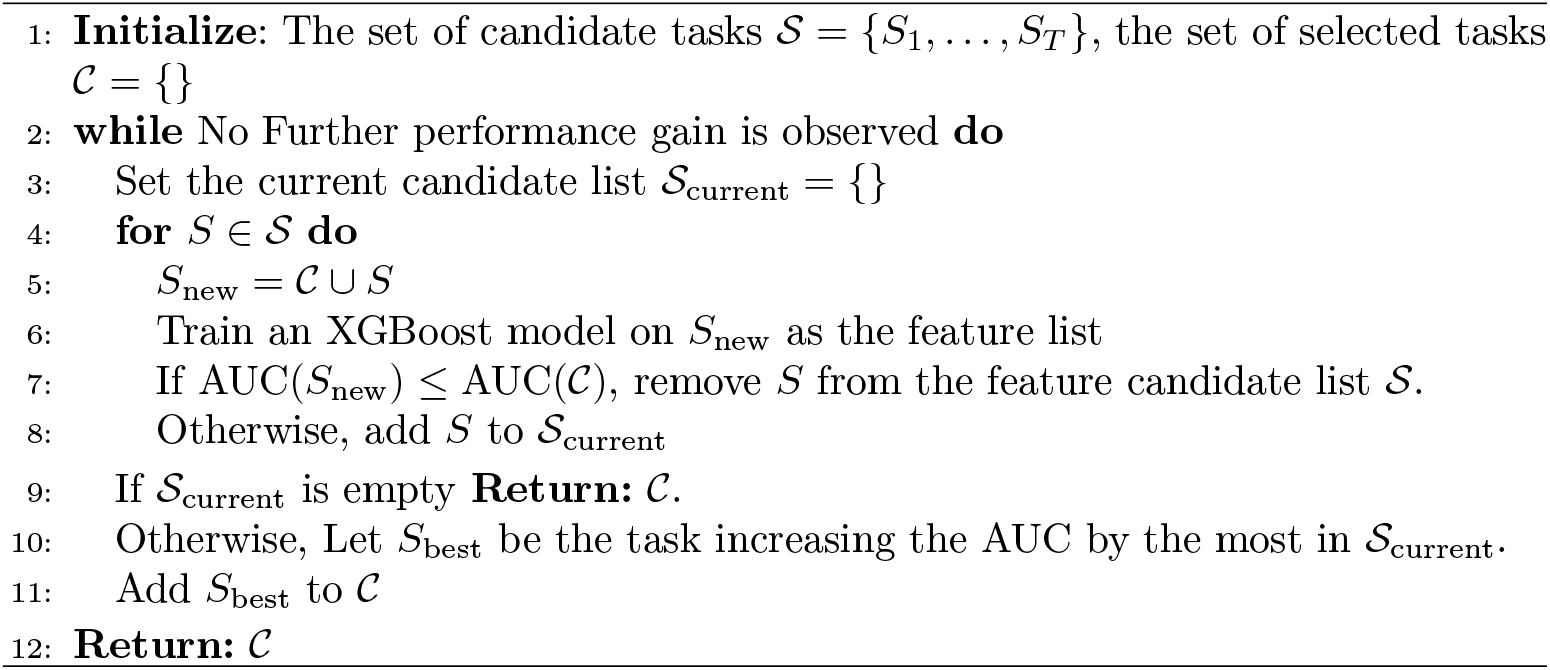

This feature selection method serves two key objectives. First, it maintains (and even slightly improves) the performance of the underlying model compared to training on the full set of features. Second, it enhances the model’s interpretability by selecting a smaller, more manageable subset of tasks and features. As a result, this approach enables researchers to focus on collecting data for these specific tasks and features, thereby reducing the complexity and cost of the data integration process for patients with Alzheimer’s Disease (AD). Figure 1 depicts the entire proposed machine learning pipeline.

## 3 Results

We compared our method to several state-of-the-art feature selection methods, including ANOVA, the Chi-square test, Fisher’s score test, correlation coefficients and the random forests feature importances as feature selection alternatives. Furthermore, we compared the performance of the developed method based on XGBoost with alternative classifiers, including Support Vector Machines, KNN, Logistic Regression, 2-layer Multi-Layer Perceptron (MLP), Decision Trees (CART implementation) and Random Forest. Our analysis shows that the proposed task and feature selection methods combined with the XGBoost classifier lead to better performance than the other approaches, while it only uses 4 tasks. Therefore, it suggests that instead of collecting data by performing a large number of tasks on patients, one can focus on these 4 tasks (Task Numbers: 8, 15, 17, and 19) to have a reliable predictor of AD. Furthermore, after selecting these tasks, we apply the feature selection method on the 4 tasks to obtain the most essential features. Figure 2 presents the selected tasks (indices in the dataset starting from 0) and their corresponding feature importances in the trained XGBoost model.

**Fig 2.**
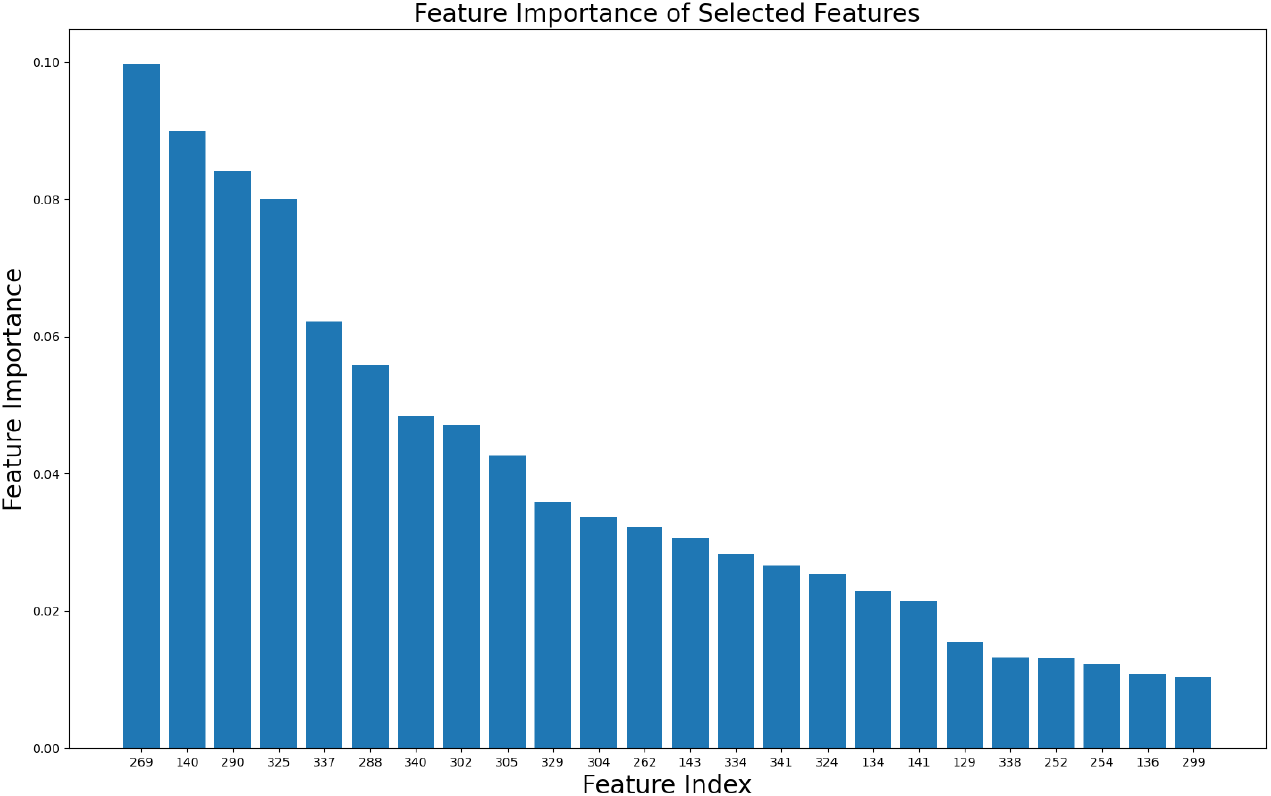
The selected features and their corresponding feature importance in the trained XGBoost model.

To evaluate the performance of these methods, we consider the Leave-One-Out Cross Validation (LOOCV) strategy as the primary performance criteria. Leave-One-Out can be seen as the K-Fold cross-validation approach where *K* is the number of folds precisely equal to *n*, the number of data points. We compute the Leave-One-Out for a given classifier 𝒞, the performance measure ℒ (e.g., AUC, Accuracy, Recall, Precision, etc.), on Dataset 𝒟 as follows:

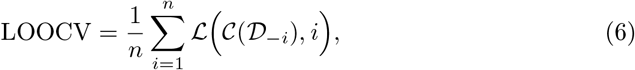

where 𝒟_−*i*_ represents the dataset 𝒟 without data point *i*. In other words, at each iteration *i*, we train the model on the dataset without data point *i* and evaluate it on the data point *i*. The LOOCV equals the average of these evaluations.

A grid search strategy optimizes the hyperparameters of all classification methods. For the feature selection part, the number of tasks and features chosen (4 and 24, respectively) are equal for a fair comparison. First, these feature selection methods are applied to the dataset as a preprocessing stage. Then, the XGBoost classifier is applied to the set of features obtained by each of these feature selection methods. Table 3 reports the performance of the proposed feature selection method and several state-of-the-art feature selection approaches with metrics, including AUC, recall, precision, and F1 score. Next, we fix our feature selection methodology and change the underlying model (both for the feature selection base method and for the downstream classification task). The results are reported in Table 4. Interestingly, the selected tasks are the same for all methods except logistic regression and SVM (Task 23 is selected instead of Task 19). The chosen task numbers for other base models are 8 15, 17, and 19.

**Table 3.** The Effect of Different Feature Selection Methods on the Performance of the XGBoost Classifier.

| Method | AUC | Recall | Precision | F1 |
| --- | --- | --- | --- | --- |
| Correlation Coefficient | 0.8944 | 90.35% | 88.46% | 0.8939 |
| Chi-Square Test | 0.8936 | 90.23% | 88.41% | 0.8931 |
| Fisher's Score Test | 0.8944 | 90.35% | 88.46% | 0.8939 |
| Random Forest Importance | 0.8994 | 90.28% | 89.41% | 0.8984 |
| XGBoost Importance | 0.9015 | 91.68% | 89.93% | 0.9079 |
| **Our Method** | **0.9137** | **92.94%** | **89.77%** | **0.9132** |

**Table 4.**
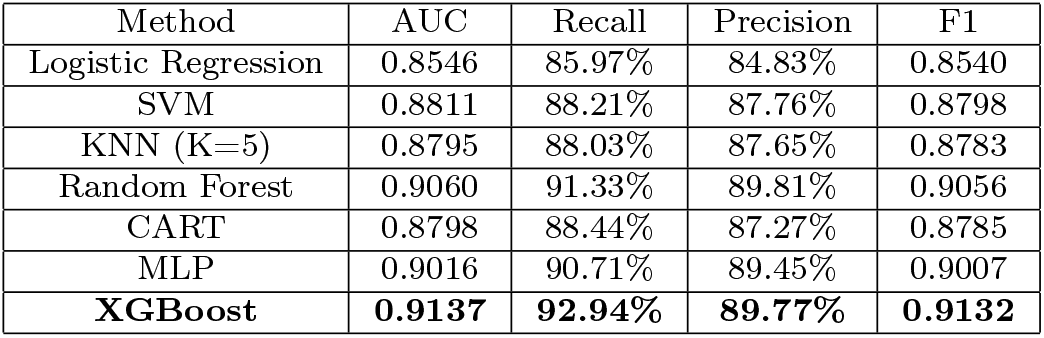
Applying Different Models as the Base Classifier for AD Detection Task.

| Method | AUC | Recall | Precision | F1 |
| --- | --- | --- | --- | --- |
| Logistic Regression | 0.8546 | 85.97% | 84.83% | 0.8540 |
| SVM | 0.8811 | 88.21% | 87.76% | 0.8798 |
| KNN (K=5) | 0.8795 | 88.03% | 87.65% | 0.8783 |
| Random Forest | 0.9060 | 91.33% | 89.81% | 0.9056 |
| CART | 0.8798 | 88.44% | 87.27% | 0.8785 |
| MLP | 0.9016 | 90.71% | 89.45% | 0.9007 |
| **XGBoost** | **0.9137** | **92.94%** | **89.77%** | **0.9132** |

## 4 Conclusion

This study presents a diagnostic methodology for AD that seeks to optimize predictive accuracy through a strategic balance of task efficiency and feature selection. By employing a two-step forward-backward feature selection process tailored to the XGBoost model, we identified a minimal yet highly informative subset of handwriting-derived features, thereby enabling superior classification performance. The model demonstrated robust metrics in accuracy, recall, precision, and F1 score, underscoring its reliability and clinical applicability. This reliability should instill confidence in its potential to improve AD diagnostics. Although our approach optimizes the balance among high predictive accuracy, interpretability, and a reduced number of tasks, some alternative models report higher accuracy. However, such models often involve trade-offs, related to increased complexity, reduced generalizability, and limited clinical applicability. Compared to other machine learning methodologies, our approach requires fewer tasks, thus reducing the burden on patients and clinicians and aligning with the need for streamlined diagnostic protocols in clinical settings. This methodology supports targeted assessments in AD diagnostics and underscores the potential of handwriting analysis as an intriguing and minimally invasive biomarker, opening up new avenues of research and application.

## Data Availability

All data referred to in the manuscript are publicly available and can be accessed from the DARWIN dataset, hosted by the UCI Machine Learning Repository

https://archive.ics.uci.edu/dataset/732/darwin?utm_source=chatgpt.com

